# Anticarcinogenic effects from plasma elderly trained: the role of mitochondrial function

**DOI:** 10.1101/2021.09.08.21263280

**Authors:** Alessandra Peres, Gilson Pires Dorneles, Gisele Branchini, Fernanda Bordignon, Pedro R T Romão, Luciele Minuzzi, Fabio S Lira, Mateus Cavalcante, Viviane Elsner

## Abstract

This study aimed to evaluate the effects of a multimodal exercise training on systemic cytokine levels of the elderly, and the impact of post-exercise training plasma on prostate cancer cell viability and proliferation *in vitro*. Fasting blood samples were collected from eight institutionalized elderly before and after eight weeks of multimodal exercise training (twice a week). The levels of interleukin(IL)-1ra, IL-1β, IL-2, IL-6, IL-10, IL-17, interferon (IFN)-α, tumor necrosis factor (TNF)-α, fibroblast growth factor (FGF)-1, platelet-derived growth factor (PDGF) and transforming growth factor (TGF)-α were evaluated in the peripheral blood. PC3 prostate cancer cell lines were incubated with 10% plasma acquired before and after exercise training from each participant. Multimodal exercise training increased the plasma levels of IL-2, IL-10, IFN-α, and FGF-1, and decreased TNF-α concentrations. PC3 cells presented decreased cell viability evaluated by MTT and lactate dehydrogenase activity as well as lower rates of cell proliferation after the incubation with post-training plasma samples. Furthermore, the incubation of PC-3 cells with post-training plasma decreased the mitochondrial membrane polarization and increased mitochondrial reactive oxygen species (ROS) production without changes in cytosolic ROS. Post-training plasma did not change apoptosis or necrosis rates in the PC-3 cell line. In conclusion, we showed that systemic adaptations in plasma mediators of institutionalized elderly might alter cell viability and proliferation by targeting mitochondrial ROS in a prostate cancer cell line.

## Introduction

Several aspects of the host immune system are impacted by biological aging, characterized by a chronic inflammatory state associated with an accumulation of senescent exhausted myeloid and lymphoid cells (1). Inflammaging is a persistent and non-resolved elevation of pro-inflammatory mediators, mainly cytokines and chemokines, in the peripheral blood of elderly individuals (2). This condition has been associated with the etiology and clinical course of most age-related diseases and mortality, including several cancer types(2). In this sense, prostate cancer is the second most common cancer in men and becomes more prevalent with aging, and inflammaging emerges as one of the main contributors to prostate carcinogenesis due to innate and adaptive inflammatory response enhancement (3).

Epidemiological data from men with non-metastatic prostate cancer suggest that regular moderate-to-vigorous physical activity in a frequency > 3 hours per week significantly reduce the risk of prostate cancer-related death (4). Furthermore, emerging preclinical and clinical studies showed mechanistic immune and physiological adaptations that may prevent or treat prostate cancer progression (5). Changes in systemic inflammation and blood factors composition induced by exercise training adaptations may directly impact cancer cell viability. Past data highlighted the role of acute exercise-conditioned serum from young and aged humans to decrease cancer cell viability (6–8). However, few studies evaluated the potential effects of exercise training to induce anticarcinogenic effects in plasma of elderly individuals(6). This study verified the impact of multimodal exercise training on systemic inflammation of institutionalized elderly. Moreover, we evaluated the role of before- and after-exercise training plasma of the elderly on viability and proliferation of an immortalized prostate cancer androgen unresponsive PC3 cells.

## Methods

### Participants

Eight institutionalized older adults (6 women, aged 73.38 ± 11.28 y, body mass index 27.8 ± 4.9 kg/m^2^, calf circumference 38.3 ± 3.6 cm) living in a long-term facility in Porto Alegre City, south of Brazil. The inclusion and exclusion criteria were previously reported (9). Participants should not be engaged in structured exercise training protocols in a period prior to 6 months before the trial. The Ethics Research Committee of Centro Universitário Metodista-IPA, Brazil approved the current study number 3.376.078). All participants signed written informed before enrollment and all procedures was in conformity with the Declaration of Helsinki.

### Study design and training protocol

Participants were submitted to a multimodal exercise training (8 weeks, 2x/week, 60 minutes each session) in this noncontrolled trial. The multimodal protocol used in the current study was based on a previous study (Marmeleira et al., 2018). Specifically, each session was divided into the four following moments: 1) warm-up (5 minutes), with stretching and active upper/lower limb exercises; 2) exercises focused on cardiovascular capacity, strength, balance/agility and flexibility (25 minutes), which included walking, stationary gait, resistance exercises for the main upper/lower muscle groups, unipodal support with open/closed arms over the chest, anterior/lateral inclination and static stretches; 3) exercises focused on perception and cognition (such as double-task), attention, memory and processing of requested actions (25 minutes), such as walking and naming fruit/colour names, completing previously established circuits, attending to requested verbal commands and memorizing motor/verbal signals; and 4) relaxation and breathing techniques (5 min).

### Blood samples and Cytokine measurement

Fasting venous blood samples were collected from the antecubital vein into EDTA tubes (8 mL), centrifuged (1000 *g*, 10 minutes), aliquoted in microtubes and stored at - 80°C. Blood collection was performed before the first exercise session and 48 h after the last exercise bout. The systemic levels of interleukin(IL)-1ra, IL-1β, IL-2, IL-6, IL-10, IL-17, interferon (IFN)-α, tumour necrosis factor (TNF)-α (all from ThermoFisher, USA), fibroblast growth factor (FGF)-1, platelet-derived growth factor (PDGF) and transforming growth factor (TGF)-α (all from RayBiotech, USA) were determined by *Enzyme Linked Immunosorbent Assay* (ELISA) following manufacturer’s instructions in a microplate reader (EzReader, Biochrom, USA).

### Cell Culture Experiments

The PC3 prostate cancer cell line from the American Type Culture Collection (ATCC® CRL-1435™) was used in this study. Cells were cultured in 75 cm^2^ flasks using Rosswell Park Memorial Institute medium-1640 (RPMI-1640) supplemented with 10% (v/v) fetal bovine serum (FBS), 0.1 mg/mL streptomycin, 100 U/mL penicillin. Cells were maintained in a humidifed incubator at 37 °C and at 5% CO2, during a maximum of 15 passages. During experiments, 10% FBS was replaced with 10% of human plasma obtained before or after the training sessions.

After a 48-h treatment, the cell viability assay (MTT and LDH release) was performed by colorimetric reduction of MTT (3-(4,5-dimethylthiazole bromide-2-yl)-2,5-diphenyltetrazolium bromide) to formazan. Samples were read by a spectrophotometer at 492 nm (10). Appropriate controls with DMSO (extract solvent) and blank liposome were performed, to eliminate the membrane solvent hydrolysis effect in results interpretation. LDH activity was evaluated in the commercial kit (LDH Roche, Brazil). Released LDH in the culture media was coupled to an enzymatic assay yielding a red color, the intensity of which was measured at 490□nm by a microplate reader. All experiments were done in triplicate and results were shown as the mean of triplicates from three independent experiments.

The proliferative response of PC3 was evaluated by the decay of CFSE fluorescence using the FACSCalibur (Becton Dickinson, San Jose, CA) flow cytometer equipped with a blue argon laser (488nm) and a 530/30nm bandpass filter. The CFSE fluorescence was analyzed in histograms of FL1 channel. The “M1 region” was defined as CFSE-stained cells derived from unstimulated cultures, which represented the peak of quiescent cells, and the M2 region was defined as proliferative cells according to the peaks of CFSE intensity. Apoptosis and necrosis of PC3 cells was measured using FITC Annexin V with propidium iodide Apoptosis/necrosis Detection Kit (556547, BD Biosciences) according to the manufacturer’s instructions using an FACSCalibur flow cytometer (BD Biosciences).

Mitochondrial membrane polarization, cytosolic and mitochondrial reactive oxygen species (ROS) analysis were performed after 12-h of *in vitro* PC3 incubation with pre- and post-exercise training plasma. The mitochondrial membrane potential (ΔΨm) was quantified according to a method previously described (11), using the fluorescent dye rhodamine 123 (Rh 123, Sigma-Aldrich, USA). Mitochondrial superoxide generation in live cells was assessed with MitoSOX Red (Invitrogen, ThermoFisher, USA). Cytosolic ROS production was evaluated using the reagent 20,70-dichlorofluorescein diacetate (DCF-DA), which becomes fluorescent when oxidated by ROS (Sigma Aldrich, USA). Analyses were performed by using CELLQuest Pro Software (BD Bioscience) on a FACSCalibur flow cytometer (BD Bioscience).

### Statistical analysis

Data were analyzed in GraphPad Prism 8.0 (USA). Data were presented as mean ± standard deviation. Before-after exercise training comparisons were verified by Paired Student t-test. p value ≤ 0.05 were considered statistically significant.

## Results

### Multimodal exercise training alters systemic cytokine levels of institutionalized elderly individuals

Increases in the plasma levels of Interleukin (IL)-2 (p=0.01), Interferon-alpha (IFN-α) (p=0.01), and Fibroblast growth factor (FGF)-1 (p=0.01) occurred after the training period. On the other hand, tumor necrosis factor-alpha (TNF-α) concentrations decreased (p=0.03) after training. In addition, IL-10 levels tended to be higher after training compared to the baseline values (p=0.06). No statistical differences were observed in IL-1ra, IL-1β, IL-6, platelet-derived growth factor (PDGF), IL-17a, or transforming growth factor-alpha (TGF-α) (p>0.05).

### Plasma acquired after multimodal exercise training alters prostate cancer cell viability and proliferation by changes in mitochondria membrane potential and increased mitochondrial reactive oxygen species production

Next, we incubated the PC3 prostate tumor cell line with 10% plasma collected before exercise training and 48 h after the exercise training period to evaluate the potential inhibitory effect of exercised plasma on cancer cell viability and growth. After 48 h of experiment, cell viability evaluated by MTT were significantly lower in PC3 cell incubated with post-training plasma (p=0.03), concomitant to higher LDH activity on the supernatant of cultured PC3 cells with post-training plasma (p=0.02). In addition, the analysis of CFSE expression showed significantly lower proliferation in post-training plasma PC3 culture combined with the pre-training plasma condition (p=0.028). Thus, multimodal exercise training induces anticarcinogenic effects in the plasma of elderly individuals by changes in cell viability and proliferation. Next, we evaluated the role of mitochondria-induced reactive oxygen species generation in the anticarcinogenic effects of plasma of trained elderly individuals. After 12-h of PC-3 cell culture, post-training plasma decreased mitochondrial membrane polarization compared to pre-training plasma incubation condition (p<0.01). The mitochondria membrane depolarization was accompanied by increased mitochondrial ROS production in the PC3 cell line incubated with post-training serum (p=0.04), without changes in cytosolic ROS generation (p>0.05). Finally, PC-3 cell line incubation with post-training plasma did not change apoptosis or necrosis cell events (p>0.05).

Post training cytokine plasma levels were correlated with *in vitro* experiments MTT, LDH, CFSE cell proliferation, cytosolic ROS, mitochondrial membrane polarization and mitochondrial ROS in PC3 prostate cancer cell line, and heat map correlation is presented in Figure 3. TNF-α levels correlated with mitochondrial ROS (r=0.82; p=0.04). IL-6 levels correlated with supernatant LDH content (r=0.94; p=0.005) and CFSE cell proliferation (r=0.82; p=0.04). IL-1ra concentrations significantly correlated with mitochondrial membrane polarization (r=0.88; p=0.01). IL-17a levels correlated with mitochondrial ROS (r=0.74; p=0.04). Finally, TGF-α inversely correlated with LDH content (r= -0.83; p=0.01).

**Figure 1.**
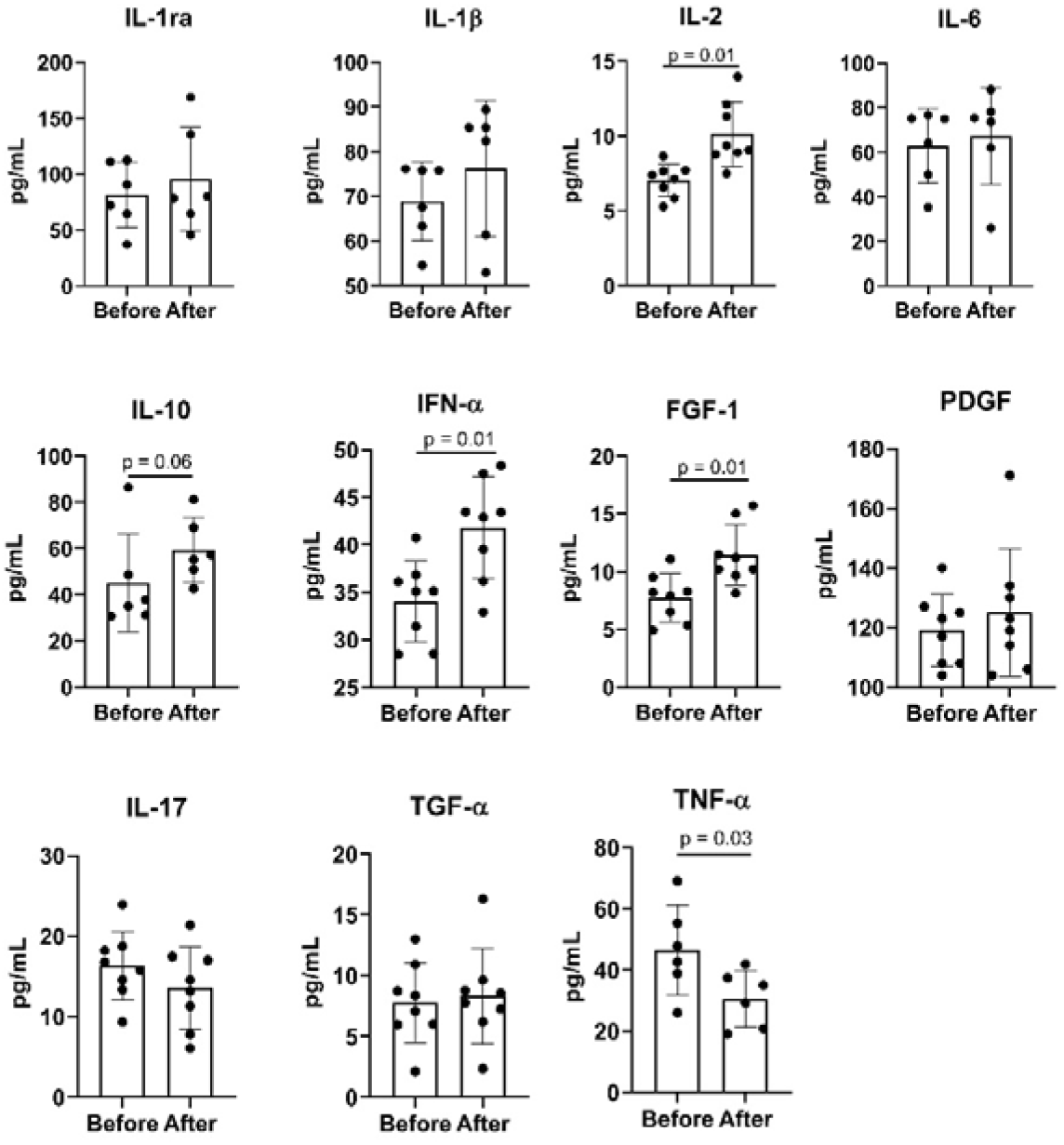
The impact of 8 weeks of multimodal exercise training on systemic cytokine levels of institutionalized elderly individuals. Statistical analysis performed by Paired t-test (p<0.05).

**Figure 2.**
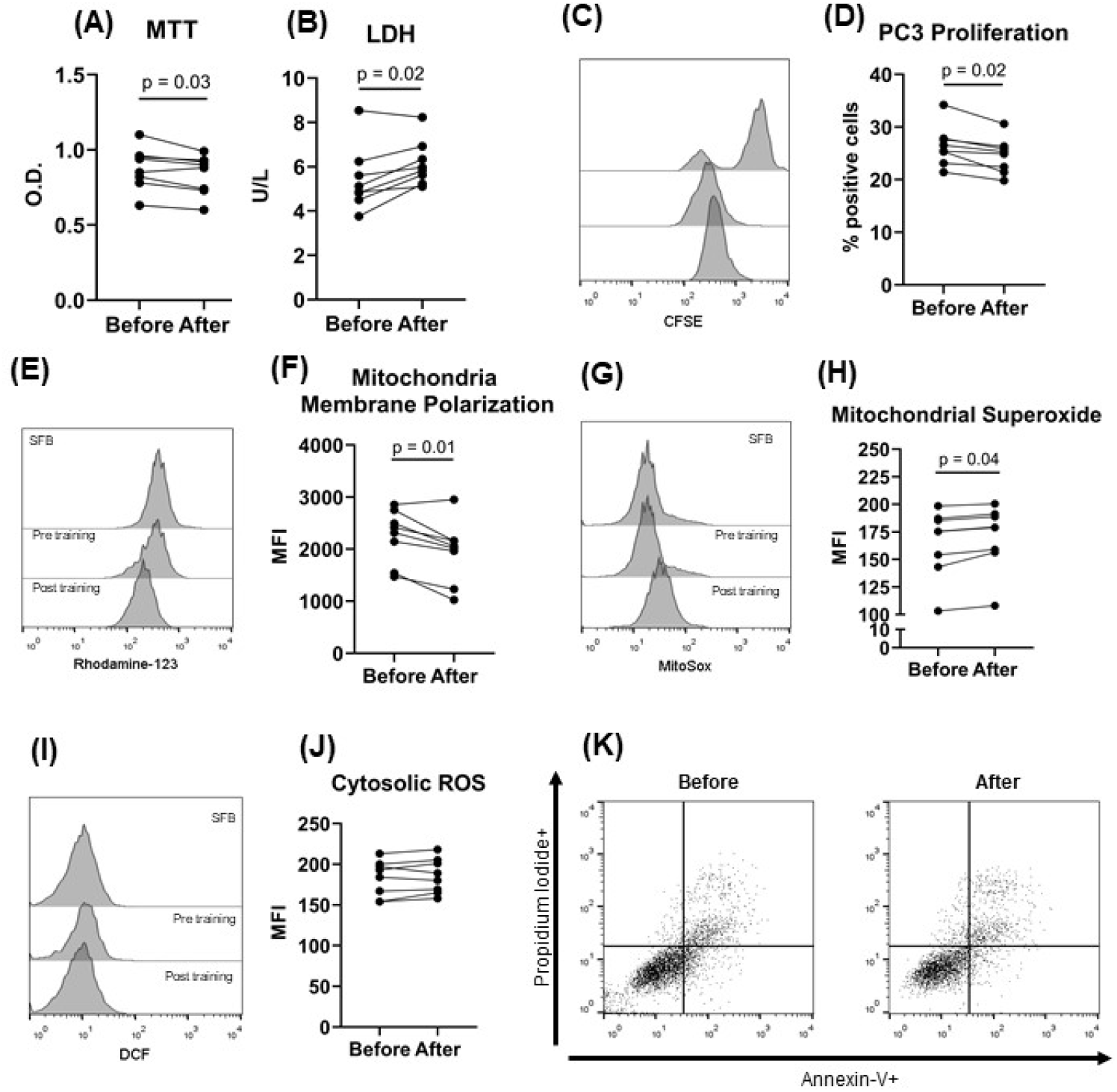
Anticarcinogenic effects of plasma from trained elderly individuals in PC3 prostate cancer cell line. PC-3 Prostate cancer cell line were incubated with plasma of institutionalized elderly individuals obtained before training and 48 h after 8 weeks of exercise training. Cell viability was evaluated by MTT (A) and LDH activity (B), and proliferation by the drop of CFSE fluorescence in PC-3 cells (C and D) after 48 h of cell culture. Histogram of rhodamine-123 (E) evaluated the mitochondrial membrane potential (F), histogram of Mitosox (G) quantified mitochondrial superoxide (H) generation, and histogram of DCF(I) evaluated cytosolic reactive oxygen species (J) production after 12-h of PC-3 cell line incubation with plasma obtained before and after exercise training. Apoptosis (Annexin-V+ cells) or necrosis (Propidium iodide+ cells) did not differ before – after comparison (K). Statistical analysis conducted through Paired student t-test (p<0.05).

**Figure 3.**
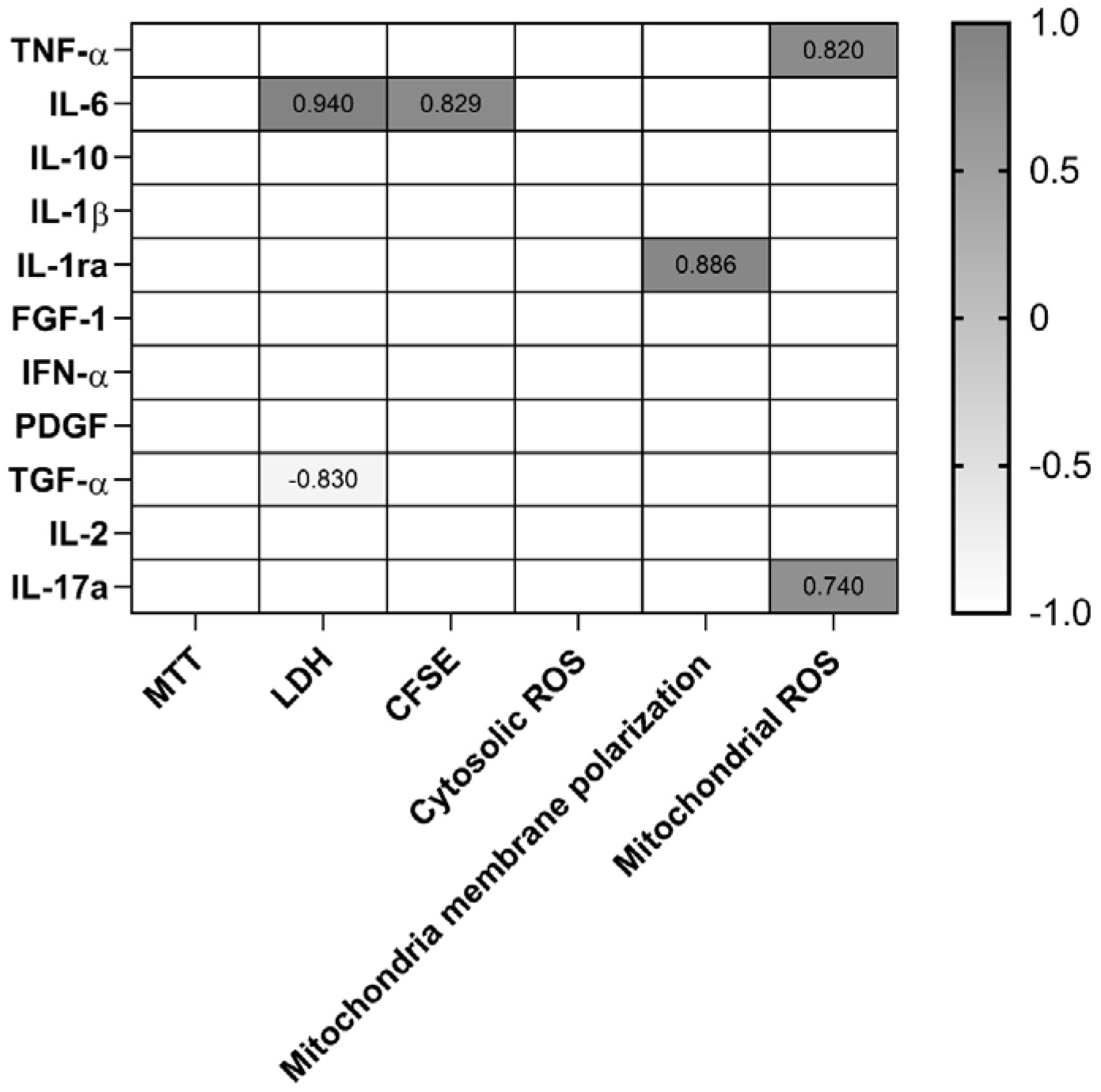
Heat map correlation between plasma cytokine levels and *in vitro* analysis of cell viability (MTT and LDH), cell proliferation (CFSE), reactive oxygen species production (cytosolic ROS and mitochondrial ROS) and mitochondria membrane polarization in PC3 prostate cancer cell line.

## Discussion

The main results of this study were: 1) 8 weeks of multimodal exercise training increases the systemic levels of IL-2, IFN-α, and FGF-1 and decreases the TNF-α concentrations in aged individuals; 2) the incubation of PC3 prostate cancer cell line with post-exercise training plasma of elderly individuals leads to lower cell viability and proliferation rates after 48 h of treatment; 3) conditioned post-training plasma-induced mitochondrial membrane depolarization and higher mitochondrial ROS, but not cytosolic ROS, in PC3 prostate cancer cells without changes in apoptosis/necrosis rate. Taken together, we showed for the first time that systemic inflammatory adaptations in blood mediators of institutionalized elderly engaged in multimodal exercise training have strong anticarcinogenic potential against prostate cancer through changes in mitochondrial membrane polarization and mitochondrial ROS generation.

Here, exercise training was able to decrease proinflammatory TNF-α levels, suggesting a role to induce anti-inflammatory profile in institutionalized elderlies. However, other classic proinflammatory mediators, such as IL-6, IL-17a and IL-1β, did not change after 8 weeks of multimodal exercise. These results may suggest that the potential anti-inflammatory adaptations observed in previous observational and longitudinal studies (12) may need a longer intervention time than 8 weeks to be achieved. On the other hand, we are the first study to observe increased FGF-1 increased after exercise training period. FGF-1, also called acidic FGF, plays an important role in the regulation of cell survival, cell division, angiogenesis, cell differentiation and migration (13). Interestingly, experimental studies shows that mice treated with FGF-1 restore blood glucose levels and endothelial function, highlighting the role of this growth factor in the vascular health and metabolic control(14). Furthermore, mutated *fgf1* gene is linked to accelerated neurological senescence profile in mice(15). Thus, FGF-1 emerges as an important biological mediator to the control of aging through exercise training.

The incubation of conditioned plasma obtained from institutionalized elderly engaged in multimodal exercise training decreased both cell viability and proliferation in PC3 prostate cancer cell line. This results are in line with the previously reported by a series of systematic reviews and meta-analysis recently published who demonstrated the anticarcinogenic potential of peripheral blood factors of exercised individuals (6,7,16). It was hypothesized that exercise-induced biochemical mediators release, mainly myokines and immunomodulatory cytokines, have a critical role in decreasing cancer cell viability and growth(5). Interestingly, apoptosis and necrosis rate were unchanged after the incubation of PC3 prostate cell line with conditioned exercised-plasma, confirming previously data who demonstrate that exercised mediators reduces cell viability without changes in cell death pathways(17,18). Notably, several studies were conducted using acute exercise session models and the tumor-suppressive effects of chronic longitudinal exercise training is poorly studied.

We found an increases in IL-2 and IFN-α levels in the peripheral blood of elderly after exercise training period. Both IL-2 and IFN-α have strong anti-tumorigenic directly effects against cancer cell, and *in vitro* cytokine treatment of prostate tumor cell lines can effectively alter a number of prostate carcinoma properties closely associated with tumor invasion and metastatic phenotype(19). In addition, the correlation between post-training cytokine levels and PC3 cell viability, proliferation and mitochondrial function revealed some associations between changes in systemic inflammatory mediators and cancer cell phenotype.

Here, we describe for the first time the mitochondrial dysfunction in PC3 prostate cell line incubated with post-exercise training plasma of elderly. Targeting cancer cell mitochondria has been long suggested as a therapeutic approach to control cell proliferation and growth. In this sense, several pharmacological therapies alter mitochondria function to induce cell death and lower tumor progression (20). In the present study we show that depolarization of the mitochondrial membrane potential associated with increasing superoxide production (mitochondrial ROS) after 12h of PC3 incubation with conditioned plasma of elderly. Furthermore, mitochondria membrane depolarization leads to translocation of apoptosis-induced factor (AIF) to the nuclei and activation of caspase-12 associated with endoplasmic reticulum to induce cell death(21). Moreover, mitochondrial membrane depolarization directly affects complex II and its function in electric chain transport, leading to ROS generation and the activation of apoptotic cascade(22). However, the lack of changes in apoptosis rate after conditioned plasma incubation may indicate the need of repeated or prolonged incubation time to induce cancer cell death.

In conclusion, this longitudinal study described for the first time the potential of conditioned plasma to decrease cell viability and proliferation in PC3 prostate tumor cell line. We also demonstrated a new mechanistic pathway by which exercise may alters prostate cell function through mitochondrial function, mainly by mitochondrial membrane depolarization and superoxide formation. These changes were accompanied by alterations in several systemic inflammatory mediators after multimodal exercise training. Collectively, changes in blood factors composition by exercise training contribute to the control of prostate tumorigenesis, suggesting the role of exercise as an adjuvant therapeutic in cancer treatment and prevention.

## Data Availability

Data will be available when requested.

## Acknowledgements

We are grateful to the Brazilian agencies Coordenação de Aperfeiçoamento de Pessoal de Nível Superior (CAPES) – Finance Code 001 for financial support. GPD is supported by postdoctoral *fellowship from* CAPES. PRTR and AP are grateful to *CNPq* for the *PQ productivity scholarship*.

